# Impact of SARS-Cov-2 on Clinical Trial Unit workforce in the United Kingdom; An observational study

**DOI:** 10.1101/2022.06.30.22277052

**Authors:** Gayathri Delanerolle, Jintong Hu, Heitor Cavalini, Lucy Yardley, Katharine Barnard-Kelly, Katheryn Elliot, Vanessa Raymont, Shanaya Rathod, Jian Qing Shi, Peter Phiri

**Affiliations:** Nuffield Department of Primary Care Health Sciences, University of Oxford, OX3 7JX, Oxford, UK; Southern University of Science and Technology, Shenzhen, 518055, China; Psychology Department, Faculty of Environmental and Life Sciences, University of Southampton, SO17 1BJ, Southampton, UK; Southern Health NHS Foundation Trust, SO40 2RZ, UK; Department of Psychiatry, University of Oxford, OX3 7JX, Oxford, UK; The Alan Turing Institute, NW1 2DB, London, UK

**Author notes:** **Corresponding author: Dr Peter Phiri, BSc, PhD, RN, Director of Research & Innovation/ Visiting Fellow,** Research & Innovation Department, Southern Health NHS Foundation Trust, Clinical Trials Facility, Tom Rudd Unit Moorgreen Hospital, Botley Road, West End, Southampton SO30 3JB, United Kingdom. Shared last author (Jian Qing Shi, Peter Phiri). Shared second author (Jintong Hu, Heitor Cavalini, Lucy Yardley). ***Declarations***. **Funding** This work is funded by the NIHR. **Availability of data and material** The authors will consider sharing the dataset gathered upon receipt of reasonable requests. **Code availability** The authors will consider sharing the dataset gathered upon receipt of reasonable requests. **Author contributions** GD and PP developed the study protocol and embedded this within the EPIC project’s work-package 3. The Chief Investigator of this study is PP and Principal Investigator GD. GD, JT and JQS designed and completed the statistical analysis. The qualitative analysis was completed by KBK, PP and GD. All authors critically appraised and commented on previous versions of the manuscript. All authors read and approved the final manuscript. **Ethics approval** HRA REC approval (21/HRA/2348) was obtained prior to the study initiation. **Consent to participate** All participants consented to take part in this study. **Consent for publication** All authors consented to publish this manuscript.

## Abstract

**Objective:** The clinical trial unit (CTU) workforce in the UK have been delivering COVID-19 research since the inception of the pandemic. Challenges associated with COVID-19 research have impacted the global healthcare communities differently. Thus, the overall objective of the study was to determine the mental health impact among CTU staff working during the COVID-19 pandemic.

**Design:** A mixed-methods based observational study was designed using a new workforce impact questionnaire using validated mental health assessments of Vancouver Index of Acculturation (VIA), Hospital Anxiety and Depression Scale (HADS), Insomnia Severity Index (ISI), Pandemic Stress Index (PSI), Burnout Assessment Too-12 (BAT-12), General Self Efficacy Scale (GSE) and The Everyday Discrimination Scale (EDS).

**Setting:** The Qualtrics platform was used to deploy the questionnaire where a quantitative analysis was conducted. The qualitative part of the study used the Microsoft Teams digital application to complete the interviews.

**Participants:** All participants were CTU staff within the United Kingdom.

## Introduction

The World Health Organization (WHO) declared the outbreak of the COVID-19 pandemic on the 11^th^ of March 2020 which impacted the clinical and clinical research workforce significantly. [1] Lockdown, using masks, social distancing and various other measures were used to manage the spread of the virus. [2] While much of the spotlight was on incidence and prevalence of COVID-19, the impact on conducting scientific research was significant. Existing clinical trials were paused especially within the UK and COVID-19 research was prioritised.[3] The clinical trial profession as a whole has a number of staffing groups with a variety of job titles within the UK. Industry, academia and the NHS struggle to recruit and retain clinical trial staff and many leave the profession. In April 2022, jobs.ac.uk indicated over 150 unfilled clinical trial positions nationally. Whilst this could be due to a variety of reasons including short term contracts, workload acumen, issues with salaries for the expected job specification and lack of flexibility for hybrid working a higher vacancy rate impacts on the existing clinical trial workforce as they would be required to cover a larger volume of work.

According to the National Institute for Health Research (NIHR), new clinical trials even for complex and urgent areas such as cancer, [5] were suspended in the UK, and staff were redeployed [6]. Clinical trials units (CTUs) are specialist units that play a key role in study design, conduct, analysis and subsequent publication. CTUs house expert clinical research staff, including clinical trialists, statisticians, epidemiologists, methodologists, quality assurance and trial management staff that have experience to setup, manage and deliver clinical trials. Some CTUs deliver a diverse array of studies, whilst others specialise in either a specific clinical area or a type of intervention such as investigational medicinal products (IMPs), medical devices and complex interventions. CTUs are legally responsible for maintaining adherence to all compliance procedures regardless of their embodiment within an academic or NHS organisation. The CTUs played an essential role during the pandemic in the UK. The Management of Health and Safety at Work Regulations (1999 as amended) that requires employers to ensure the work environment is, as far as reasonably practicable, safe and without risks to health. The COVID-19 pandemic introduced an unforeseen risk that had not been considered previously.

Some countries have reported challenges to manage their research during the pandemic, facing challenges what include physical violence perpetrated towards members of the medical staff,[8] unreliable therapies, [3] and prejudice amongst ethnic minorities health workers in frontlines against de virus.[9] However, the impact on the CTU workforce has not been explored. Hence, this study explores the COVID-19 impact with an aim to report the challenges and develop strategies to better develop pandemic frameworks in the future.

## Methods

The CTU workforce in the UK is approximated around 25,000 although some staff have shared roles with multiple departments spanning across academic and NHS organisations. We designed a mixed methods observational-digital study to explore the experiences of CTU staff within the UK. The survey was deployed digitally via the NIHR, UKTMN, social media and the UK CRC.

### Aims

The primary aim of the study was to determine the mental health impact due to the challenges endeavoured by the clinical trial workforce whilst delivering clinical research during the COVID-19 pandemic. The secondary aim of the study was to determine the impact on the wellbeing on staff that would aid to propose a suitable pandemic preparedness framework for clinical trial units and staff conducting studies during a pandemic.

### Eligibility criteria

All participants that were 18 years and above, employed within a CTU in the UK were included in the study. Participants also required to have access to a smartphone, tablet, or computer to complete the survey online, and willing to provide informed e-consent.

### Data collection and extraction

Quantitative data was collected through an online questionnaire using the Qualtrics XM platform. The sample size comprised of 485 participants.

A subgroup of the sample was randomly invited to take part in semi-structured interviews. All interviews were conducted via a secure online facility (password protected teams teleconference), audio-recorded and transcribed in full. Data collection and analysis was integrated with a process based on framework methodology used to analyse the data including development of a coding frame based on identified key themes and detailed coding of transcripts.

### Outcome measures

Quantitative measures were used to evaluate mental health impact byway of validated questionnaires. For the purpose of this study, we used Vancouver Index of Acculturation (VIA), Hospital Anxiety and Depression Scale (HADS), Insomnia Severity Index (ISI), Pandemic Stress Index (PSI), Burnout Assessment Too-12 (BAT-12), General Self Efficacy Scale (GSE) and The Everyday Discrimination Scale (EDS). Cut-off scores were used to ensure the erroneous decisions for each mental health assessment completed by all study participants could be unified for the purpose of the analysis.

[Table 9]

### Analysis plan

#### Quantitative

The analysis focused on 9 questionnaires with 14 dimensions. Participants were divided into several subgroups based on age, gender, ethnicity, role and length of service. Mean and standard deviation scores on 14 dimensions were calculated. ANOVA and t-tests were applied to check the difference in means between different subgroups.

A correlation heat-map was used to demonstrate the correlation between each questionnaire based on Spearman correlation. An item-total correlation was calculated to focus on the core items. Total item correlation in this instance refers to the correlation coefficient summarised between each specific item and other items, which is the quantification of the importance of specific item.

Linear regression was used to investigate the mental health impact during the pandemic.

#### Qualitative

Transcripts were analysed using thematic and content analysis. Two experienced qualitative researchers independently reviewed transcripts and conduct analyses. A coding framework was developed to capture key themes with each coded theme subjected to detailed analyses to identify subthemes and illustrative quotes.

## Results

### Quantitative

A sample of 485 patients completed the questionnaire. Key characteristics of the sample are indicated in Table 1 compromising of all sexes, ages and geographies. Of 485, 257 did not disclose their professional category and job title, whilst 60 and 228 participants reported these, respectively.

### Age

Of 485 respondents, more than half (50.7%) were aged between 35-54 years. The sample included 2.3% of young adults aged between 18-24 years. Cognitive impairment, one of the four dimensions measured by BAT-12, showed significant differences between age groups. People aged between 18-24 and 35-54 years scored higher compared with other groups. The young adult group scored the highest scores for cognitive impairment, depression and daily discrimination. The elderly group, aged over 65 years of age had the highest scores for heritage and mainstream, GSE and lowest everyday discrimination score. T-tests showed anxiety level of white British were significantly lower than of ethnic minorities. People aged 45-54 years have the highest anxiety with a score of 11 or over (proportion of 45.1%), while people aged 25-34 years have the lowest anxiety proportion of 39.6%. The anxiety proportion for people aged 18-24, 25-34, 35-44, 45-54, 55-64, 65 years and over were 40.0%, 39.6%, 40.0%, 45.1%, 44.4%, and 43.8%, respectively. However, the p-value (0.8092) indicated the anxiety level is independent with length of service.

### Gender

Approximately 73.4% of the participants were female and 15.5% were male. Anxiety level was measured by anxiety score, which is the summation of the odds items within the HADS questionnaire. Male participants had a lower averaged anxiety score of 9.2 in comparison to female, 10.4. Of the total 356 female respondents, 237 completed the anxiety questionnaire, thus there was a 33.4% of missingness. Of the 237 women, 110 (46.4%) had an anxiety score of 11 or higher and, 92 (38.8%) reported an anxiety score of 8 to 10. In comparison, of the total 75 male respondents, 48 completed anxiety questionnaires. Of the 48 participants, 9 (18.8%) had an anxiety score of 11 or higher and, 29 (60.4%) had a score of 8-10. The Chi-square test indicated a p-value of 0.001808 suggesting levels of anxiety appears to be similar across the genders.

Men scored significantly lower than women on the dimension of exhaustion (7.8 and 8.7, respectively). Women appear to be less influenced by mainstream and heritage cultures in comparison to men as per the low scores observed from the VIA questionnaire.

In terms of ethnicity and race, 70.1% of the respondents were white British and 30.9% were of ethnic minorities of African, Asian, Bangladeshi and Indian. Ethnic minorities were groups for the purpose of conducting a meaningful analysis to evaluate their mental health outcomes.

[Table 1, Table 2, Table11, and Table 12]

### Mental health assessment

#### Length of service

The BAT-12 scores appear to increase with the length of service as indicated in Table 2. Thus, elevated exposure to exhaustion, mental distance, cognitive and emotional impairment appear to be common observations within this group. The ANOVA test showed statistically significant scores between cognitive impairment and mental distance among different age groups (less than 1 year, 1 to 5 years, 6 to ten years, 11 to 15 years, 16 to 20 years, over 21 years) with p-values of 0.008 and less than 0.001 respectively. P-values were not significant within the exhaustion and emotional impairment dimensions with 0.329 and 0.363, respectively. A trend analysis indicates the overall exhaustion and emotional impairment levels increased with the length of service ranges from less than a year to 20 years. There appears to be a large variation within the data. We excluded the data for the group of over 21 years since the sample size is rather small and it is treated as an outlier.

[Table 2]

#### Well-being

The pooled data indicated 25.8% of respondents suffered from long-term conditions and 3.9% from disabilities. Participants (28.2%) who had an existing mental health condition prior to the pandemic, believed their disease did not worsen, thus a lack of a notable change in their wellbeing was reported. Approximately, 10.9% of participants with mental health conditions felt better since the pandemic began. Participants that were mental health naive (43.1%) their wellbeing worsened since the pandemic began. In 36.3% of participants, physical health conditions did not change since the beginning of the pandemic began whilst 13.4% of participants felt there were improvements. Approximately 32.8% of participants reported their physical condition worsened since the pandemic began.

[Table 1]

Of the trial management participants, 28.3% suffered from long-term conditions which is slightly higher. Clinical trials reported their mental health and physical health conditions worsened since the pandemic began, in comparison to other professional groups with 49.6% and 35.4%, respectively. These participants also the use of medications for their mental and physical conditions; 15.7% and 25.2%, respectively.

In comparison, the remaining participants had a lower percentage of medication use for mental and physical health with 12.6% and 19.8, respectively. The cognitive impairment level is relatively high with an average impairment score of 8.40 among the clinical trialist group compared with the rest of study participants (7.93).

[Table 1 and Table 5]

#### Professional group outcomes

Five different professional groups with acceptable sample size (ranging from 8 to 127, mean 46.4) were considered into analysis (trial management, quality assurance, database management, doctor/nurse, statistician). Statisticians had the highest level of exhaustion, mental distance, and depression whilst Quality assurance staff had the highest level of cognitive impairment. Nurses and doctors appear to have the highest anxiety and emotional impairment. An overall statistical significance cannot be determined based on the professional group sample sizes. Although, this has clinical significance in relation to the wellbeing of staff. The demographic characteristics and its association with burnout, HADS, everyday discrimination, ISI and GSE is indicated in Table 2.

Approximately, 28.3% of participants working in trial management suffered from long-term conditions and is slightly higher than that of the study population. On the other hand, clinical trialists felt their mental and physical health had worsened since the pandemic began in comparison to other professional groups, 49.6% and 35.4%, respectively.

[Table 5]

#### Heat-map correlation

Based on the heatmap, heritage and mainstream scores were highly associated with each other based on the spearman correlation score of 0.8. Anxiety level showed a strong correlation with levels of depression, insomnia, exhaustion, emotional impairment and cognitive impairment where the spearman correlation greater than 0.4. GSE and everyday discrimination showed a correlation with the four dimensions of BAT-12 although the spearman correlation was less than -0.4 and over 0.3, respectively.

[Figure 1]

#### Linear regression

To investigate whether long term exposure to high level stressful environment has influence to their mental health, we combined anxiety, depression and burn out scores (denoted by *y*) and plot it against the length of service (using the mid-point for each group, denoted by *x*) in Figure 2. We found that the overall score is increasing along the length of service except the group of over 21.

#### Item-total correlations of ISI

Means and standard deviations for the ISI and sleep diary measures are reported in Table 3. The ISI average total score was 7.8 8 (SD 5.86). The internal consistency of the ISI was estimated with a Cronbach alpha coefficient and by the item-total correlations. The internal consistency, (i.e. degree of consistency or homogeneity of the items within a scale) of the ISI was 0.74. The item-total correlations varied from a low of 0.55 (initial) to a high of 0.78 (interference) with an average of 0.70 (see Table 3).

#### Geographical

Of the 485 respondents, 137 disclosed their longitude and latitude, accounting for about 59%. Vast majority of respondents were from England, concentrated to the central and southern parts of England, including Southampton, Bristol, London, Oxford and Birmingham. Scotland had the second largest number of respondents primarily from Edinburgh, Glasgow and Dundee.

[Table 10]

#### Qualitative

For the qualitative part of the study, 6 participants (4 women and 2 men) consented and completed interviews (supplementary document). Interview duration ranged from twenty-six to fifty-four minutes. Interview participants included a senior statistician, research nurses and trials managers. Participants described their work-related stress levels between a 3 and a 6. Two participants felt things were worse since the beginning of the COVID-19 pandemic, whilst one said there have been improvements and the other claimed things had returned to pre-COVID-19 levels. There was a general sense that the job came with periods of high pressure that created stress, however this was unrelated to COVID-19.

Participants reported pre-COVID-19 related daily work issues associated with the clinical trials they were working on, office environment and the commute. Different views were reported on the perceived effectiveness of open plan offices in terms of noise, distractions and challenges associated with some processes required for clinical trials management.

There were also tasks that could no longer be completed as before due to changes to the workforce availability. Research nurses transferred to support COVID-19 patients meant recruitment had to be halted for most studies and others transitioned to an ‘on-line’ or ‘telephone’ mode of delivery where possible

Views diverged on contributors to stress, for example,

‘*now you have a bit more spare time and are not being pestered by colleagues all of the time on the office, it helps’ [participant 002]; compared to ‘things went very scary, very high for every area of life for everybody …. This is how we operate now, there’s certainly an acceptance so not very high but the bar is set a lot lower now’ [participant 003]. One participant stated that they needed a certain level of stress to be effective ‘I reckon if my work-related stress was below a five, I probably wouldn’t have any impetus to do anything’ [participant 004]*.

At the height of COVID, however, there was an overwhelming sense of things being beyond individual control for example;

‘*it was awful…*.. *in terms of your day, your day wasn’t your own … the adrenalin kicked in because everyone was going into survival mode …’ [003]. Similarly, ‘mostly things that are out of my control …*.. *things that are out of my hand that are stressful … such as recruitment levels, trying to get that sorted’ [001]; ‘… the whole situation was intensely stressful’ [006]*.

Participants reported challenges with balancing personal and professional that caused additional stress. Lack of dedicated or appropriate workspace was problematic, as was sharing dining table space with partners who were also trying to work from home. Similarly, one participant was having building works done which had been suspended due to COVID-19, thus had only one usable room in the house for long periods of time during the first lockdown. One participant’s husband worked in A&E and said;

‘*had a hell of a time …. He’s lost so many people’ [005]*

Another participant’s husband worked away and was doing so due to COVID. During the initial lockdown she was unable to get a childcare place for her five year-old son *‘so I was home-schooling on my own, trying to work at home. That was the most stressful point I think because it’s virtually impossible with a five year-old to home school and work at the same time, so I was working evenings and weekends and it was just constant and then, because we were in lockdown we didn’t see anyone, we didn’t go anywhere, we were totally isolated*., *So it was pretty awful*.*’ [006]*.

There was a consistent theme of the sudden disappearance of a traditional ‘working day’ where working extended often unsociable hours. This change was perceived differently by the participants;

‘… *so nobody had an escape plan, nobody had down time but 10, 11 o’clock at night emailing isn’t ideal’ [003]. ‘I respond to emails any time eg at 11pm I can give a quick answer and it’s done. It’s good and bad. I’m quite responsive with people, I’m not a stickler with hours. [004]. ‘There is an expectation to work more from one person who wants to have meetings at 6-7 in the evening. As such as lockdown was done I didn’t want to do that anymore’ [002]*.

The transition from face-to-face meetings to online zoom meetings came with its own technical challenges including frequent back-to-back meetings without a break or IT failures.

‘*I used to have two meetings a week on zoom and there was a time when there were 7 a day*’ [003]. ‘*You bounce from meeting to meeting rather than doing it physically’ [001]. There were positives, however with one participant saying ‘I do think your true self comes out, everyone’s much more relaxed in a zoom environment than we were before because you have to be. That’s the norm and it’s taken the pressure off*.’ [003].

The hybrid approach to work also changed the future working requirements.

‘*There aren’t many people showing up to work in the office, it feels pointless to go in’ [001]. ‘It was quite hard because it’s a big open plan office which is why we’re not sort of back in the office yet because they’re trying to work out ventilation and hot desking and stuff…… a key part of what helps me do my work is a big pair of noise-cancelling headphones because it’s too loud and I get distracted really easily’ [004]. ‘Technically we’re back but not everyone is yet. All the desks have moved. Having to travel again is weird. There are no face-to-face meetings, just getting back to being back close to people on the commute again’. [005] There were also positive experiences however ‘I didn’t realise till I started to go back in just how refreshing it is to have, like, even a laugh with someone in your office. Just to have that two minutes’ [005]*

Communication became very difficult for some participants;

‘*The biggest problem was in the communication …. We realised no-one’s coming back full-time and had to adapt to different ways of working and communicating’ [005]*.

*‘In the early days, the doors were just closed. That was it, just closed’ [003]*.

One participant felt there was added workload in terms of administration ‘there’s a lot more applications we have to fill in; ….

*‘Basically you’ve got to apply to work, to use anywhere and get approval, use the clinic rooms for studies’ [006]*.

Overall, there was sense of uncertainty despite the pandemic status in the UK changing. There were mixed views on the desirability of office working and returning to a pre-COVID-19 working style.

## Discussion

Based on the collective findings are evident of a negative mental health impact on the CTU workforce due to the COVID-19 pandemic. There are varying degrees of experiences of stress and burnout when comparing those with and without a mental illness or a physical condition. A clinical review would be beneficial for these patients.

COVID-19 has demonstrated the importance of diversity and the disproportionate impact minority populations endeavour. Based on our study participant pool, there appears to be under representation of ethnic minorities. Adequate representation could have allowed us to have more meaningful conclusions about their impact of COVID-19 on their mental health.

The mental health impact identified may result in increased sickness absences. This would also have a negative impact on performance and efficiency. Whilst this study cannot determine the precise medium and long-term effects of increased sickness absences at an individual and organisational level, a negative impact can be determined if the overall wellbeing remains the same.

It appears, there is significant variability in experiences and opinions around *lived experience* professionally and personally during the pandemic. Participants reflected powerful insights into the impact of COVID-19 whilst working to deliver COVID-19 research in a CTU environment.

## Limitations

Enrolment of participants could have been increased if there was less pressure on the CTU workforce. Cultural adaptations could have been considered when conducting further work based on the evidence gathered within this study if there ethnic minorities were better represented.

There were considerable challenges to increase participants for the qualitative component of the study. Longitudinal data collection could be a useful step to continue to assess these findings that could aid employees and employers alike to make quality improvements.

## Conclusion

Our study indicates the substantial personal impact the CTU workforce has endeavoured during the pandemic. Viability of sustainable clinical trial conduct is based on multi-professional involvement in CTU settings thus, it would be in the interest of all healthcare professionals to improve the support systems available to better manage working conditions especially as part of pandemic preparedness.

Recommendations to inform Future Preparedness in supporting the CTU workforce in delivering pandemic and non-pandemic research include ensuring continuity and clarity of communication via different media, providing opportunities for flexibility in working hours, within reasonable constraints to ensure staff are not pressured to work during times they would not usually do so. Providing opportunities for collaborative problem-solving via different media as per the needs of individual team member. Maintaining consistency, where possible, in frequency and content of contact so that staff feel supported in different aspects of their working roles. Contingency planning supplies of necessary equipment for effective home working, with consideration of all relevant health and safety legislation to ensure the wellbeing of staff.

## Data Availability

All data produced in the present study are available upon reasonable request to the authors

## Acknowledgements

The authors acknowledge support from Southern Health NHS Foundation Trust.

