## Supplementary material for "Impact of SARS-Cov-2 on Clinical Trial Unit workforce in the United Kingdom; An observational study": Tables and figures

***Table 1. Demographic information of all participants***

| **Demographics** | **Questionnaire respondents (N = 485)** | | |
| --- | --- | --- | --- |
| **Variables** | **N(%)** | **Variables** | **N(%)** |
| Age | 18 - 24 11 (2.3%)  25 - 34 90 (18.6%)  35 - 44 131 (27.0%)  45 - 54 115 (23.7%)  55 - 64 70 (14.4%)  65 and over 21 (4.3%)  Not available 47 (9.7%) | Religion | Christian 135 (27.8%)  Muslim 9 (1.9%)  Hindu 1 (0.2%)  Jewish 1 (0.2%)  Buddhist 2 (0.4%)  Sikh 2 (0.4%)  No religion 268 (55.3%)  Other 13 (2.7%)  Not available 54 (11.1%) |
| Gender | Female 356 (73.4%)  Male 75 (15.5%)  Other 2 (0.4%)  Prefer not to say 5 (1.0%)  Not available 47 (9.7%) | Ethnic | African 1 (0.2%)  Asian 8 (1.6%)  Bangladesh 2 (0.4%)  Black 2 (0.4%)  Indian 5 (1.0%)  Pakistani 4 (0.8%)  White & Asian 3 (0.6%)  White & Black African 3 (0.6%)  White & Caribbean 2 (0.4%)  White British 340 (70.1%)  White Irish 3 (0.6%)  Other background 34 (7.0%)  Prefer not to say 6 (0.6%)  Not available 72 (14.8%) |
| Nationality | United Kingdom 265 (54.6%)  Not available 220 (45.4%) |  |  |
| Healthcare professional | Yes 60 (12.4%)  No 379 (78.1%)  Not available 46 (9.5%) |  |  |
| Job title | Administration / Management 193 (39.8%)  Attending / Staff physician 2 (0.4%)  Physician assistant / Nurse practitioner 3 (0.6%)  Registered nurse 25 (5.1%)  Unit assistant / Clerk / Secretory 5 (1.0%)  Other 208 (42.9%)  Not available 49 (10.1%) | | |
| Professional group (Healthcare professional = Yes  N = 60) | Academic research 5 (8.3%)  Allied Health Professional 4 (6.7%)  Allied Health Professional by training 4 (6.7%)  Clinical Trialist 5 (8.3%)  Doctor 4 (6.7%)  Nurse 31 (51.7)  Other 6 (10.0%) | | |
| **Health and wellbeing** | | | |
| **Suffer from any long-term conditions** | Yes 125 (25.8%)  No 277 (57.1%)  Prefer not to say 7 (1.4%)  Not available 76 (15.7%) | Suffer from any disabilities | Yes 19 (3.9%)  No 384 (79.1%)  Prefer not to say 3 (0.6%)  Not available 79 (16.3%) |
| **Taking medication for any mental health condition** | Yes 65 (13.4%)  No 337 (69.5%)  Prefer not to say 6 (1.2%)  Not available 77 (15.9%) | Taking medication for any physical condition | Yes 103 (21.2%)  No 297 (61.2%)  Prefer not to say 5 (1.0%)  Not available 80 (16.5%) |
| **Mental health rating since the pandemic began** | Much better 15 (3.1%)  Somewhat better 38 (7.8%)  About the same 137 (28.2%)  Somewhat worse 173 (35.7%)  Much worse 36 (7.4%)  Not available 86 (17.7%) | Physical health rating since the pandemic began | Much better 19 (3.9%)  Somewhat better 46 (9.5%)  About the same 176 (36.3%)  Somewhat worse 141 (29.1%)  Much worse 18 (3.7%)  Not available 85 (17.5%) |
| **Test positive for COVID-19 in the past 12 months** | Yes 51 (10.5%)  No 350 (72.2%)  Not available 84 (17.3%) |  |  |

***Table 2.*** ***Demographic characteristics with burnout, HADS, everyday discrimination, ISI and GSE***

|  |  | **BAT-12** | | | | **HADS** | | **Everyday discrimination** | **ISI** | **GSE** | **VIA** | |
| --- | --- | --- | --- | --- | --- | --- | --- | --- | --- | --- | --- | --- |
| **Variable** | N | Exhaustion | Mental distance | Cognitive impairment | Emotional Impairment | Anxiety | Depression |  |  |  | Heritage | Mainstream |
| **Gender** | | | | | | | | | | | | |
| Female | 356 | 8.7 (2.7) | 7.3 (2.9) | 8.1 (2.4) | 5.5 (2.4) | 10.4 (2.6) | 8.4 (1.7) | 0.8 (0.7) | 8.1 (5.8) | 3.0 (0.5) | 6.44 (1.47) | 6.60 (1.44) |
| Male | 75 | 7.8 (2.7) | 7.5 (2.6) | 7.9 (2.8) | 5.2 (2.1) | 9.2 (2.5) | 8.4 (1.5) | 0.8 (0.6) | 6.7 (6.1) | 3.1 (0.4) | 5.99 (1.46) | 6.20 (1.56) |
| T-test |  | 0.041* | 0.607 | 0.667 | 0.445 | 0.002* | 0.986 | 0.965 | 0.149 | 0.548 | 0.060* | 0.092* |
| **Age** | | | | | | | | | | | | |
| 18 - 24 | 11 | 10.2 (4.3) | 7.7 (3.3) | 8.3 (3.3) | 5.7 (2.4) | 10.4 (2.7) | 9.5 (2.9) | 0.9 (0.7) | 10.2 (8.1) | 2.8 (0.6) | 6.21 (1.05) | 6.09 (1.58) |
| 25 - 34 | 90 | 8.7 (2.8) | 7.2 (2.8) | 7.7 (2.5) | 5.4 (2.5) | 9.8 (2.4) | 8.2 (1.6) | 0.7 (0.7) | 7.7 (6.6) | 3.1 (0.4) | 6.38 (1.49) | 6.65 (1.27) |
| 35 - 44 | 131 | 8.6 (2.7) | 7.6 (3.0) | 8.6 (2.6) | 5.4 (2.4) | 9.7 (3.2) | 8.1 (1.7) | 0.8 (0.7) | 6.8 (5.7) | 3.0 (0.5) | 6.31 (1.46) | 6.44 (1.44) |
| 45 - 54 | 115 | 8.8 (2.6) | 7.8 (2.8) | 8.2 (2.5) | 5.9 (2.7) | 10.4 (2.7) | 8.6 (1.6) | 0.9 (0.7) | 8.6 (5.5) | 3.0 (0.5) | 6.27 (1.45) | 6.41 (1.53) |
| 55 - 64 | 70 | 7.8 (2.7) | 6.9 (2.8) | 7.3 (2.2) | 5.0 (2.0) | 9.8 (2.4) | 8.9 (1.6) | 0.6 (0.6) | 8.2 (5.6) | 3.1 (0.4) | 6.38 (1.67) | 6.64 (1.54) |
| > 64 | 21 | 8.1 (3.2) | 5.7 (2.2) | 7.1 (2.4) | 5.2 (1.7) | 9.7 (3.2) | 9.0 (1.6) | 0.5 (0.4) | 8.6 (6.2) | 3.3 (0.4) | 7.51 (1.37) | 7.80 (1.47) |
| ANOVA |  | 0.276 | 0.147 | 0.042* | 0.521 | 0.699 | 0.016* | 0.054* | 0.376 | 0.070* | 0.213 | 0.089* |
| **Ethnicity** | | | | | | | | | | | | |
| White British | 340 | 8.5 (2.6) | 7.3 (2.8) | 8.2 (2.4) | 5.4 (2.3) | 10.1 (2.5) | 8.5 (1.7) | 0.7 (0.6) | 7.9 (5.9) | 3.0 (0.4) | 6.25 (1.50) | 6.52 (1.49) |
| Other ethnicity | 67 | 8.8 (3.0) | 7.2 (3.1) | 7.8 (2.8) | 5.7 (2.8) | 10.9 (2.8) | 8.2 (1.7) | 0.9 (0.8) | 8.4 (5.8) | 3.0 (0.6) | 6.78 (1.30) | 6.59 (1.29) |
| T-test |  | 0.562 | 0.806 | 0.403 | 0.460 | 0.078* | 0.193 | 0.223 | 0.638 | 0.570 | 0.020* | 0.751 |
| **Length of service** | | | | | | | | | | | | |
| Less than 1 year | 50 | 7.7 (2.9) | 5.8 (2.3) | 7.0 (2.5) | 4.7 (2.1) | 9.7 (2.3) | 8.4 (1.8) | 0.7 (0.6) | 7.7 (6.6) | 3.1 (0.4) | 6.36 (1.81) | 6.39 (1.76) |
| 1 to 5 years | 192 | 8.7 (2.7) | 7.9 (2.8) | 8.2 (2.3) | 5.6 (2.5) | 10.2 (2.7) | 8.4 (1.7) | 0.8 (0.7) | 7.9 (5.9) | 3.0 (0.4) | 6.35 (1.36) | 6.57 (1.34) |
| 6 to 10 years | 105 | 8.7 (2.7) | 7.3 (2.9) | 8.3 (2.7) | 5.6 (2.5) | 10.7 (2.7) | 8.5 (1.6) | 0.8 (0.7) | 7.8 (5.6) | 3.1 (0.5) | 6.37 (1.56) | 6.52 (1.58) |
| 11 to 15 years | 47 | 8.8 (2.9) | 7.2 (2.7) | 8.1 (2.6) | 5.3 (2.6) | 9.8 (2.0) | 8.4 (1.9) | 0.7 (0.5) | 8.3 (6.2) | 3.1 (0.6) | 6.17 (1.41) | 6.23 (1.42) |
| 16 to 20 years | 7 | 9.3 (1.3) | 8.0 (1.4) | 7.8 (2.1) | 6.3 (0.6) | 12.2 (3.6) | 8.6 (0.9) | 0.8 (0.2) | 6.6 (2.9) | 3.0 (0.2) | 7.12 (2.00) | 7.62 (1.43) |
| Over 21 years | 10 | 6.6 (3.8) | 3.4 (0.9) | 4.6 (1.8) | 4.0 (1.4) | 8.8 (2.7) | 9.5 (0.8) | 0.4 (0.4) | 9.3 (6.8) | 3.0 (0.3) | 6.72 (1.59) | 6.76 (1.57) |
| ANOVA |  | 0.329 | <0.001* | 0.008* | 0.363 | 0.106 | 0.744 | 0.690 | 0.978 | 0.830 | 0.852 | 0.504 |
| **Professional groups** | | | | | | | | | | | | |
| Trial management | 127 | 8.4 (2.5) | 7.4 (2.8) | 8.4 (2.8) | 5.4 (2.3) | 7.9 (4.3) | 4.8 (3.6) | 0.8 (0.7) | 7.1 (5.5) | 3.1 (0.5) |  |  |
| Quality assurance | 8 | 9.0 (1.9) | 7.1 (1.8) | 8.7 (2.0) | 4.4 (1.3) | 8.7 (4.6) | 3.5 (4.0) | 0.9 (0.6) | 10.3 (8.3) | 3.0 (0.2) |  |  |
| Database management | 34 | 8.5 (3.0) | 7.2 (3.0) | 8.3 (2.3) | 6.1 (2.7) | 8.0 (3.9) | 5.3 (4.0) | 0.7 (0.6) | 9.1 (6.9) | 2.8 (0.6) |  |  |
| Doctor/Nurse | 34 | 8.8 (3.1) | 7.1 (2.9) | 7.7 (2.6) | 6.2 (3.0) | 8.9 (4.2) | 5.9 (4.3) | 0.9 (0.8) | 8.5 (5.7) | 3.0 (0.3) |  |  |
| Statistician | 29 | 9.2 (3.1) | 7.8 (2.8) | 7.8 (2.5) | 5.4 (2.7) | 8.6 (4.4) | 6.1 (4.7) | 0.5 (0.7) | 6.5 (5.2) | 3.1 (0.4) |  |  |
| ANOVA |  | 0.851 | 0.965 | 0.723 | 0.364 | 0.820 | 0.480 | 0.409 | 0.368 | 0.209 |  |  |

*Note: Significant values(with p-value less than 0.1) are marked red*

***Table 3. Means (SD), item-total correlations for the ISI***

| **Severity** | **Mean (SD)** | **Item-total r** |
| --- | --- | --- |
| Initial (Difficulty falling asleep) sleep onset | 0.99 (1.10) | 0.5456 |
| Middle (Difficulty staying asleep) sleep maintenance | 1.18 (1.12) | 0.7461 |
| Terminal (Problem waking up too early) | 1.16 (1.19) | 0.6095 |
| Satisfaction | 1.87 (1.20) | 0.7554 |
| Interference | 1.08 (1.02) | 0.7758 |
| Noticeability | 0.90 (1.00) | 0.7232 |
| Distress (Worried) | 0.70 (0.84) | 0.7579 |
| Total | 7.88 (5.86) |  |

***Table 4. Summarized table of scale information***

| **Scale information** | | | |
| --- | --- | --- | --- |
| **Vancouver Index of Acculturation (VIA)** | | | |
| Heritage score | 6.36 / 9 (1.48) | Mainstream score | 6.53 / 9 (1.47) |
| **Hospital Anxiety and Depression Scale (HADS)** | | | |
| Anxiety score | 10.24 / 21 (2.60) | Depression score | 8.46 / 21 (1.69) |
| **Insomnia Severity Index (ISI)** | | | |
| Insomnia score | 7.88 / 28 (5.86) |  |  |
| **Pandemic Stress Index (PSI)** | | | |
| Behavioral experiences during COVID-19 | No changes to my life or behaviour  Practicing social distancing  Isolating or quarantine yourself  Caring for someone at home  Working from home  Not working  A change in use of healthcare services  Following media coverage related to COVID-19  Changing travel plans | | 3 (0.62%)  278 (57.32%)  154 (31.75%)  43 (8.87%)  268 (55.26%)  8 (1.65%)  107 (22.06%)  233 (48.04%)  207 (42.68%) |
| **Impact of COVID-19 on day-to-day life** | Not at all  A little  Much  Very Much  Extremely | | 4 (0.82%)  50 (10.31%)  66 (13.61%)  93 (19.18%)  65 (13.40%) |
| Physical and mental experiences during COVID-19 | Being diagnosed with COVID-19  Fear of getting COVID-19  Fear of giving COVID-19 to someone else  Worrying about friends, family, partners, etc.  Stigma or discrimination from other people  Personal financial loss  Frustration or boredom  Not having enough basic supplies  More anxiety  More depression  More sleep, less sleep, or other changes to your normal sleep pattern  Increased alcohol or other substance use  A change in sexual activity  Loneliness  Confusion about what COVID-19 is, how to prevent it, or why social distancing/isolation/quarantines are needed  Feeling that I was contributing to the greater good by preventing myself or others from getting COVID-19  Getting emotional or social support from family, friends, partners, a counsellor, or someone else  Getting financial support from family, friends, partners, an organisation, or someone else  Other difficulties or challenges | | 40 (8.25%)  135 (27.84%)  185 (38.14%)  236 (48.66%)  23 (4.74%)  41 (8.45%)  160 (32.99%)  24 (4.95%)  151 (31.13%)  70 (14.43%)  115 (23.71%)  79 (16.29%)  43 (8.87%)  106 (21.86%)  24 (4.95%)  167 (34.43%)  89 (18.35%)  10 (2.06%)  49 (10.10%) |
| **General Self Efficacy Scale (GSE)** | | | |
| Average self efficacy score | 3.04 / 4 (0.46) |  |  |
| **Burnout Assessment Tool-12 (BAT-12)** | | | |
| Exhaustion score | 8.57 / 15 (2.75) | Cognitive impairment score | 8.05 / 15 (2.51) |
| Mental distance  score | 7.33 / 15 (2.86) | Emotional impairment score | 5.46 / 15 (2.42) |
| **The Everyday Discrimination Scale (EDS)** | | | |
| Average discrimination score | 0.77 / 5 (0.65) |  |  |

***Table 5. Demographic information of participants working in trial management***

| **Demographics** | **Questionnaire respondents (N = 127)** | | |
| --- | --- | --- | --- |
| **Variables** | **N(%)** | **Variables** | **N(%)** |
| Age | 18 - 24 1 (0.8%)  25 - 34 23 (18.1%)  35 - 44 46 (36.2%)  45 - 54 35 (27.6%)  55 - 64 17 (13.4%)  65 and over 5 (3.9%) | Religion | Christian 33 (26.0%)  Muslim 7 (5.5%)  Hindu 1 (0.8%)  Buddhist 1 (0.8%)  No religion 83 (65.4%)  Other 1 (0.8%)  Not available 1 (0.8%) |
| Gender | Female 105 (82.7%)  Male 22 (17.3%) | Ethnic | Asian 2 (1.6%)  Indian 1 (0.8%)  Bangladeshi 2 (1.6%)  Pakistani 2 (1.6%)  White & Asian 1 (0.8%)  White & Caribbean 1 (0.8%)  White British 100 (78.7%)  Other background 11 (8.7%)  Not available 7 (5.5%) |
| Nationality | United Kingdom 77 (60.6%)  Not available 50 (39.4%) |  |  |
| **Health and wellbeing** | | | |
| **Suffer from any long-term conditions** | Yes 36 (28.3%)  No 83 (65.4%)  Prefer not to say 1 (0.8%)  Not available 7 (5.5%) | Suffer from any disabilities | Yes 5 (3.9%)  No 113 (89.0%)  Prefer not to say 2 (1.6%)  Not available 7 (5.5%) |
| **Taking medication for any mental health condition** | Yes 20 (15.7%)  No 100 (78.7%)  Not available 7 (5.5%) | Taking medication for any physical condition | Yes 32 (25.2%)  No 86 (67.7%)  Prefer not to say 2 (1.6%)  Not available 7 (5.5%) |
| **Mental health rating since the pandemic began** | Much better 2 (1.6%)  Somewhat better 9 (7.1%)  About the same 45 (35.4%)  Somewhat worse 54 (42.5%)  Much worse 9 (7.1%)  Not available 8 (6.3%) | Physical health rating since the pandemic began | Much better 3 (2.4%)  Somewhat better 13 (10.2%)  About the same 58 (45.7%)  Somewhat worse 39 (30.7%)  Much worse 6 (4.7%)  Not available 8 (6.3%) |
| **Test positive for COVID-19 in the past 12 months** | Yes 12 (9.4%)  No 107 (78.7%)  Not available 8 (6.3%) |  |  |

***Table 6. Demographic information of participants working in quality assurance***

| **Demographics** | **Questionnaire respondents (N = 8)** | | |
| --- | --- | --- | --- |
| **Variables** | **N(%)** | **Variables** | **N(%)** |
| Age | 25 - 34 2 (25.0%)  35 - 44 2 (25.0%)  45 - 54 3 (37.5%)  55 - 64 1 (12.5%) | Religion | Christian 3 (37.5%)  No religion 5 (62.5%) |
|  |  | Gender | Female 6 (75.0%)  Male 2 (25.0%) |
| Nationality | United Kingdom 7 (87.5%)  Not available 1 (12.5%) | Ethnic | White British 8 (100.0%) |
| **Health and wellbeing** | | | |
| **Suffer from any long-term conditions** | Yes 2 (25.0%)  No 6 (75.0%) | Suffer from any disabilities | No 8 (100.0%) |
| **Taking medication for any mental health condition** | Yes 1 (12.5%)  No 7 (87.5%) | Taking medication for any physical condition | Yes 2 (25.0%)  No 6 (75.0%) |
| **Mental health rating since the pandemic began** | Somewhat better 1 (12.5%)  About the same 3 (37.5%)  Somewhat worse 2 (25.0%)  Much worse 2 (25.0%) | Physical health rating since the pandemic began | Somewhat better 2 (25.0%)  About the same 5 (62.5%)  Somewhat worse 1 (12.5%) |
| **Test positive for COVID-19 in the past 12 months** | No 8 (100.0%) |  |  |

***Figure 1. Heatmap of scale scores(based on Spearman correlation)***

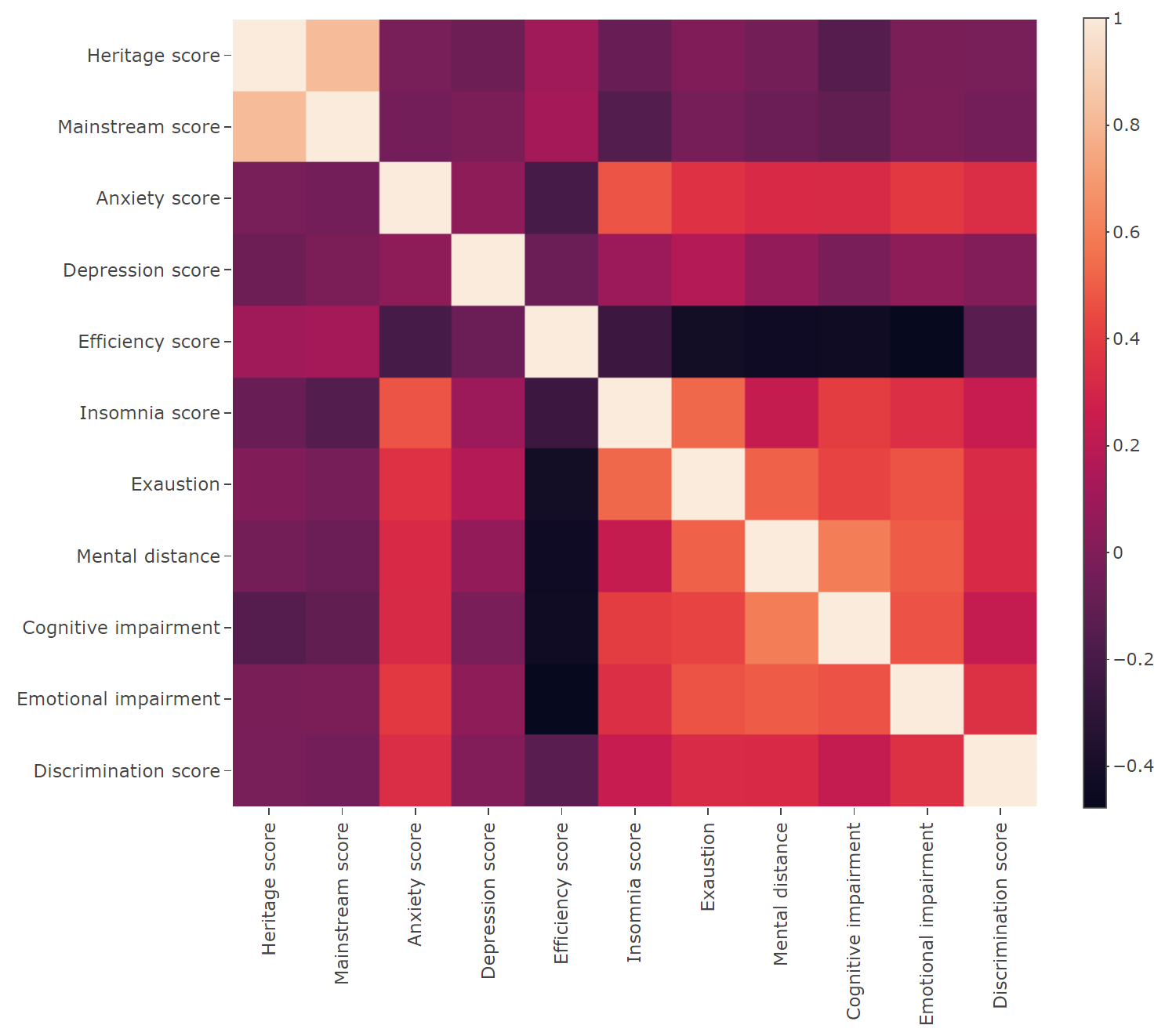

***Table 7. Professional groups with burnout, HADS, everyday discrimination, ISI and GSE***

|  |  | **BAT-12** | | | | **HADS** | | **Everyday discrimination** | **ISI** | **GSE** |
| --- | --- | --- | --- | --- | --- | --- | --- | --- | --- | --- |
| **Professional groups** | N | Exhaustion | Mental distance | Cognitive impairment | Emotional Impairment | Anxiety | Depression |  |  |  |
| Trial management | 127 | 8.4 (2.5) | 7.4 (2.8) | 8.4 (2.8) | 5.4 (2.3) | 7.9 (4.3) | 4.8 (3.6) | 0.8 (0.7) | 7.1 (5.5) | 3.1 (0.5) |
| Quality assurance | 8 | 9.0 (1.9) | 7.1 (1.8) | 8.7 (2.0) | 4.4 (1.3) | 8.7 (4.6) | 3.5 (4.0) | 0.9 (0.6) | 10.3 (8.3) | 3.0 (0.2) |
| Database management | 34 | 8.5 (3.0) | 7.2 (3.0) | 8.3 (2.3) | 6.1 (2.7) | 8.0 (3.9) | 5.3 (4.0) | 0.7 (0.6) | 9.1 (6.9) | 2.8 (0.6) |
| Doctor/Nurse | 34 | 8.8 (3.1) | 7.1 (2.9) | 7.7 (2.6) | 6.2 (3.0) | 8.9 (4.2) | 5.9 (4.3) | 0.9 (0.8) | 8.5 (5.7) | 3.0 (0.3) |
| Statistician | 29 | 9.2 (3.1) | 7.8 (2.8) | 7.8 (2.5) | 5.4 (2.7) | 8.6 (4.4) | 6.1 (4.7) | 0.5 (0.7) | 6.5 (5.2) | 3.1 (0.4) |
| ANOVA |  | 0.851 | 0.965 | 0.723 | 0.364 | 0.820 | 0.480 | 0.409 | 0.368 | 0.209 |

***Table 8. Table of Questionnaires***

| ***Questionnaire*** | ***Cut-off scores*** | ***Construct*** | ***Analytic Rationale*** | ***Dimensions*** |
| --- | --- | --- | --- | --- |
| Demographic information | None | Equity issues | Moderator | One dimension |
| Hospital Anxiety and Depression Scale (HADS) | A total score of 11 or higher indicates the probable presence of the mood disorders with a score of 8 to 10 being just suggestive of the presence of the respective state. | Psychological impact - Anxiety & Depression | Outcome measure | Anxiety score (odds items)  Depression score (even items) |
| General Self-Efficacy (GSE) | None | Assessment of general self- efficacy | Moderator | One dimension |
| Pandemic Stress Index (PSI) | None | Psychological impact | Outcome measure | One dimension |
| Insomnia Severity Index (ISI) | A score of: 0-7 is indicative of no insomnia;  8-14 indicative of a sub-threshhold insomnia;  15-21 moderate insomnia;  22-28 is indicative of severe insomnia; | Psychological impact - Sleep Quality | Outcome measure | One dimension |
| Vancouver Index of Acculturation (VIA) | None | Disadvantages and equity issues – cultural context | Moderator or mediator | Heritage score  Mainstream score |
| Burnout Assessment Tool (BAT-12) | None | Psychological impact | Moderator | Exhaustion score  Mental distance score  Cognitive impairment score  Emotional impairment score |
| The Everyday Discrimination Scale (EDS) | None | Workplace and occupational | Moderator or mediator | One dimension |
| AHRQ Patient safety culture survey | None | Workforce and occupational | Moderator or mediator | One dimension |

***Table 9. Means (SD) and ANOVA test of individual items on the EDS between different ethnicity***

| **Items of EDS** | **White British (n = 340)** | **Other (n = 67)** | **Prefer not to say (n = 6)** | **ANOVA** |
| --- | --- | --- | --- | --- |
| Treated with less courtesy | 1.3 (1.2) | 1.6 (1.5) | 1.8 (1.5) | 0.221 |
| Treated with less respect | 1.3 (1.2) | 1.5 (1.5) | 1.8 (1.5) | 0.446 |
| Receive worse service | 0.5 (0.8) | 0.7 (1.0) | 0.8 (0.8) | 0.273 |
| People act as though they think you are not intelligent | 1.1 (1.2) | 1.1 (1.3) | 0.6 (0.6) | 0.653 |
| People act as though they are afraid of you | 0.4 (0.8) | 0.8 (1.3) | 1.4 (1.7) | 0.015* |
| People act as though they think you are dishonest | 0.3 (0.6) | 0.4 (0.7) | 1.0 (0.7) | 0.054* |
| People act as though they are better than youd | 1.4 (1.2) | 1.5 (1.4) | 1.6 (1.5) | 0.792 |
| Call your names or insults you | 0.3 (0.7) | 0.5 (0.8) | 0.4 (0.9) | 0.314 |
| Threatened or assaulted | 0.1 (0.5) | 0.3 (0.6) | 0.0 (0.0) | 0.262 |

***Table 10. Geographical information of professional groups***

|  |  | **Professional groups** | | | | |
| --- | --- | --- | --- | --- | --- | --- |
| **Country** | **City** | **Trial management** | **Quality assurance** | **Nurse/Doctor** | **Database management** | **Statistician** |
| England | Plymouth | 1 | 1 |  |  |  |
|  | Southampton | 4 | 1 | 2 |  |  |
|  | Portsmouth |  |  |  |  |  |
|  | Bristol | 5 |  |  |  | 1 |
|  | London | 12 |  | 3 | 2 | 1 |
|  | Oxford | 5 |  |  | 4 | 1 |
|  | Cambridge |  |  |  | 1 | 1 |
|  | Birmingham | 7 | 1 | 1 |  |  |
|  | Norwich |  |  |  | 2 | 1 |
|  | Leicester | 1 |  |  | 3 |  |
|  | Nottingham | 1 |  |  | 1 | 1 |
|  | Chester | 2 |  |  |  |  |
|  | Liverpool | 4 |  |  | 1 | 1 |
|  | Manchester |  |  |  |  | 1 |
|  | Sheffield | 1 |  |  |  |  |
|  | Leeds |  |  |  |  |  |
|  | York | 3 |  |  |  |  |
|  | Newcastle | 1 |  |  |  |  |
|  | Colebrooke | 1 |  |  |  |  |
|  | Liss Forest | 1 |  |  |  |  |
|  | Lydiard Millicent | 1 |  |  |  |  |
|  | Aston Tirrold | 1 |  |  |  |  |
|  | Woodley | 1 |  |  |  |  |
|  | Marlow | 2 |  |  |  |  |
|  | Eton | 1 |  |  |  |  |
|  | Bicester | 1 |  |  |  |  |
|  | Aylesbury | 1 |  |  |  |  |
|  | Houghton Regis | 1 |  |  |  |  |
|  | Harlow | 1 |  |  |  |  |
|  | Stoke-on-trent | 2 |  |  |  |  |
|  | Guildford |  | 1 |  |  |  |
|  | Dawley |  | 1 |  |  |  |
|  | Arreton |  |  | 1 |  |  |
|  | Hull |  |  | 1 |  |  |
|  | Brighton |  |  |  | 2 |  |
|  | Chinnor |  |  |  | 1 |  |
|  | Manea |  |  |  |  | 1 |
| Welsh | Cardiff | 2 |  |  | 1 | 1 |
|  | Swansea |  |  |  |  |  |
|  | Newport |  | 1 |  |  |  |
|  | Aberoynon | 1 |  |  |  |  |
|  | Cwmbran |  |  |  | 2 |  |
| Scotland | Edinburgh | 4 |  |  | 2 |  |
|  | Glasgow |  | 1 | 8 |  |  |
|  | Aberdeen | 1 |  |  | 1 |  |
|  | Dundee | 3 |  | 2 |  | 1 |
|  | Inverness | 1 |  |  |  |  |
|  | Cleghorn | 1 |  |  |  |  |
|  | Broxburn | 1 |  |  |  |  |
|  | Memsie |  |  | 1 |  |  |
| Northern Ireland | Belfast | 1 |  |  |  |  |
|  | Omagh | 1 |  |  |  |  |
| Total | | 77 | 7 | 19 | 23 | 11 |

***~~Note: “Other” refers to the small and less-known cities.~~***
